# Patient Reported Tolerance of 90 Days of Pre-Operative Endocrine Therapy: Results from the POWER Trial

**DOI:** 10.1101/2025.01.28.25321286

**Authors:** Trish Millard, Lena Turkheimer, Jenna Schlefman, Gina Petroni, David Brighton, Shayna Showalter

## Abstract

**Background:** Prospective randomized data supports radiation omission in women ≥ 65 years who take adjuvant endocrine therapy (AET) following breast-conserving surgery. Many patients who omit radiation stop AET early due to side effects. In the POWER trial, a prospective single-arm study, patients took 90 days of pre-operative endocrine therapy (pre-ET) to assess tolerance before making adjuvant treatment decisions. We hypothesized that patient-reported outcomes (PROs) during pre-ET would be heterogeneous and that 90 days was sufficient time for symptoms to develop.

**Patients and Methods:** PRO data from POWER trial participants was obtained before, during, and after pre-ET, including health-related quality of life (HRQoL), depression, and ET symptoms using the EORTC-QLQ, CESD-R, and BCPT-SCL tools. PRO assessments were further analyzed after stratifying patients by high or low perceived sensitivity to medicine (PSM).

**Results:** Pre-ET PROs were assessed for 75 participants. The majority (73.3%) reported symptoms during pre-ET. Only 10.7% had symptoms severe enough to stop pre-ET before 90 days. Vasomotor (42.7%) and musculoskeletal (41.3%) symptoms were the most common. HRQoL was preserved for 66.6% participants. Patients with high PSM had more ET side effects.

**Conclusion:** Patients developed similar side effects during pre-ET as those typically seen with AET. PROs and the impact of pre-ET on HRQoL were patient-dependent. A 90-day course of pre-ET is sufficient for patients to develop symptoms reflective of long-term AET. Future analyses will assess the association of pre-ET PROs with AET initiation and adherence.

## Introduction

Historically, early-stage, estrogen receptor-positive (ER+) breast cancer has been treated with breast-conserving surgery (BCS) followed by radiation therapy (RT) and adjuvant endocrine therapy (AET). CALGB 9343 and PRIME II were practice-changing prospective randomized trials that support radiation omission for older patients. In the CALGB 9343 trial, patients ≥70 years with node-negative ER+ breast tumors 2cm or less were randomized to RT followed by AET or to AET alone.^1^ Similarly, in the PRIME II trial, patients aged ≥ 65 years with node-negative ER+ breast tumors up to 3cm were randomized to BCS followed by AET with or without RT.^2^ Long-term data from the CALGB 9343 and PRIME II trials demonstrate a modest increase in local recurrence but no difference in overall survival. In accordance with this data, the National Comprehensive Cancer Network guidelines were updated in 2004 and again in 2024, stating that RT omission is appropriate in patients ≥ 65 years with tumors ≤ 3 cm who are node-negative and plan to take AET.^3^ Despite this, the majority of this population is still treated with RT, raising concern for over-treatment.^4,5^ Decisions about adjuvant treatment are inherently complex. Many physicians still view RT omission as substandard care. Additionally, RT is often recommended due to poor adherence to AET in this population, specifically outside of a clinical trial setting.^6,7^ Pursuing RT helps to avoid potential under-treatment and worse oncologic outcomes if a patient does not adhere to AET.

Despite recommendations for AET, non-adherence is reported in 23-28% of patients in large clinical trials and is even higher (32-73% based on real-world data).^5–7^ Premature discontinuation of AET is often due to medication side effects. In older women, musculoskeletal symptoms are most commonly cited as the reason for discontinuing AET.^8^ Patient tolerance to AET is nuanced and influenced by their preferences, emphasis on health-related quality of life (HRQoL), and comorbidities. When formulating adjuvant treatment plans, patients and physicians must consider the individual patient’s likelihood of completing at least five years of AET.

The **P**re**-O**perative **W**indow of **E**ndocrine Therapy to Inform **R**adiation Therapy Decisions (POWER) trial was designed to assess whether 90 days of pre-operative endocrine therapy (pre-ET) influenced patient preferences and physician recommendations for adjuvant RT in older women with ER+ breast cancer who were candidates for RT omission (NCT04272801). While side effects from AET are common, it is not possible to predict the incidence and severity of symptoms and a patient’s willingness to continue on AET while experiencing them. Models designed to predict AET nonadherence based on patient demographics and co-morbidities perform poorly and are not clinically useful.^9^ While there is evidence that AET side effects develop as early as 90 days into the adjuvant treatment course, no studies specifically focus on symptoms and patient-reported outcomes (PROs) within a 90-day course of pre-ET.^8,10^ Additionally, it is unknown if symptoms that occur during pre-ET will ultimately be indicative of long-term adherence to AET.

This study aimed to determine if patients enrolled in the POWER trial developed typical AET side effects, measured by PROs, during the 90 days of pre-ET and whether this negatively impacted their HRQoL. Relatedly, we sought to determine if patients’ perceived sensitivity to medicine (PSM) correlated with their self-reported side effects to pre-ET. We hypothesized that a subset of patients would develop side effects in the 90-day pre-ET period and that patients who viewed themselves as sensitive to medicine would have more side effects from pre-ET.

## Methods

### The POWER Trial

Women aged ≥ 65 years with ER+, HER2 negative tumors ≤ 2 cm with clinically normal axillary lymph nodes were eligible for the POWER trial. Patients with prior ET treatment or a previous ipsilateral breast RT history were excluded. Participants were treated with 90 days of pre-ET (tamoxifen or an aromatase inhibitor (AI)). Patients were encouraged to complete the pre-ET course, but those who elected to stop early remained in the study and proceeded with BCS. All others underwent BCS following the prescribed course of pre-ET. Adjuvant decisions for RT and AET were left to the discretion of the treating physicians and patients. The primary endpoint for the POWER trial was the change in physician and patient radiation preference, which will be reported in a future analysis.

### PRO Questionnaires

Patient symptom burden and HRQoL were measured through validated PRO surveys at baseline, day 30, and day 90 of pre-ET. HRQoL was assessed using the EORTC Quality of Life Questionnaire (QLQ) C30, which consists of 30 questions on a Likert scale^11^. EORTC QLQ-C30 includes specific functional scales (physical functioning, role functioning, emotional functioning, cognitive functioning, and social functioning) and HRQoL. All scales range from 0 to 100, with a higher score representing a higher/improved functional level for HRQoL. Depression was assessed using the Center for Epidemiologic Studies Depression Scale Revised (CESD-R), which is a 20-question survey scored 0 to 60, with higher scores indicating more depression symptomatology and a score above 16 indicating risk of clinical depression.^12^ The Breast Cancer Prevention Trial Symptom Checklist (BCPT-SCL) is an 18-question survey scored on a Likert scale of 0-4 to measure general symptom burden and ET side effects.^13^ It assesses the following symptom clusters: vasomotor instability, musculoskeletal symptoms, cognitive problems, weight gain or body image concerns, vulvovaginal problems, and bladder control issues. It is scored by averaging individual item scores (range 0-4), and higher scores indicate worse symptoms. Participants completed the Perceived Sensitivity to Medicine scale at baseline, which included five questions on a Likert Scale (1 - 5) to assess if they believed they were sensitive to medications.^14^ There is a maximum score of 25, with high scores indicating high perceived sensitivity.

### Statistical Analysis

The POWER trial was powered for the primary endpoint to detect a predetermined change in physician and patient RT preference. This secondary analysis is descriptive of PRO scores at baseline and after 90 days of pre-ET. Analyses were performed to examine the mean change from baseline to day 90 in HRQoL, depression, and symptom burden clusters. Further analysis was conducted on the patients who had a decrease in HRQoL by EORTC-QLQ C30, an increase in symptom burden for specified symptom clusters measured by BCPT-SCL, and an increase in depressive symptoms measured by CESD. PRO assessments were analyzed after stratifying patients into two groups based on their patient-reported PSM. Patients were considered to have high PSM if their score was 17 or greater and low PSM if their score was less than 17. Graphical displays were used to show the observed cumulative rates of subjects with changes in their pre- and post-ET PRO scores.

## Results

### Patient Characteristics

Eighty-four patients signed informed consent to participate in the POWER Trial. One was ineligible, and four patients withdrew consent and had no follow-up information. Four additional patients were deemed unevaluable due to failure to complete essential survey components, leaving 75 patients in this analysis. The majority of participants were white (90.7%) with a mean age of 73 (range 65-92). Most patients took an AI (85.3%) (Table 1).

**Table 1.** Baseline Patient Characteristics.

| **Characteristics** | **Patients (N = 75)** |
| --- | --- |
| **Age** |  |
| Median (range) – years | 73 (65 – 92) |
| **Race – n (%)** |  |
| Black or African American | 7 (9.3%) |
| White | 68 (90.7%) |
| **Ethnicity – n (%)** |  |
| Hispanic or Latino | 1 (1.3%) |
| Non-Hispanic | 74 (98.7%) |
| **ECOG performance status score – n (%)** |  |
| 0 | 52 (69.3%) |
| 1 | 20 (26.7%) |
| 2 | 3 (4.0%) |
| **BMI** |  |
| Mean (SD) | 30.37 (7.00) |
| **Comorbidities – n (%)** |  |
| Cardiovascular Disease | 8 (10.7%) |
| Hypertension | 47 (62.7%) |
| Diabetes | 15 (20.0%) |
| Lung Disease | 8 (10.7%) |
| Osteoporosis/Osteopenia | 28 (37.3%) |
| Arthritis | 29 (38.7%) |
| Headache/Migraine | 4 (5.3%) |
| Deep Vein Thrombosis/Pulmonary embolism | 3 (4.0%) |
| Cognitive Impairment/Dementia | 1 (1.3%) |
| Depression | 14 (18.7%) |
| Smoking | 29 (38.7%) |
| **Hormonal Therapy – n (%)** |  |
| Anastrozole | 16 (21.3%) |
| Letrozole | 48 (64.0%) |
| Tamoxifen | 11 (14.7%) |
| **Primary Tumor Classification – n (%)** |  |
| T1a | 9 (12.0%) |
| T1b | 38 (50.7%) |
| T1c | 28 (37.3%) |
| **Histologic Type – n (%)** |  |
| IDC | 58 (77.3%) |
| ILC | 11 (14.7%) |
| Mucinous | 1 (1.3%) |
| Other | 5 (6.7%) |
| **Histologic grade – n (%)** |  |
| 1 | 38 (50.7%) |
| 2 | 36 (48.0%) |
| 3 | 1 (1.3%) |
| **Progesterone receptor status – n (%)** |  |
| Positive | 66 (88.0%) |

### Patient Reported Outcome Measures: HRQoL, Depression, General Symptom Burden

Between day 1 and 90, 33.3% of patients had a decrease in HRQoL (mean change -17.3), 45.3% had no change in HRQoL, and 21.3% had an improvement in HRQoL (mean change 20.8). Depression increased from day 1 to 90 in 16% of patients. Most participants (73.3%) experienced an increase in any breast cancer or ET symptom (by BCPT-SCL) while taking pre-ET. The most common symptoms reported were vasomotor and musculoskeletal symptoms. Vasomotor symptoms (i.e., hot flashes and night sweats) increased for 42.7% of patients with a median change of 1.0. and 41.3% experienced worsening musculoskeletal symptoms (i.e., general aches and pains, joint pain, and muscle stiffness) with a median increase of 0.7. Weight gain or worsening body image was reported by 22.7% of participants, with a median change of 0.5. Additionally, 33.3% of participants reported worsening bladder control (median change 1.0), 22.7% reported worsening cognitive impairment (median change 0.7), and 10.7% reported vulvovaginal problems (median change 0.5) (Figure 1).

**Figure 1:**
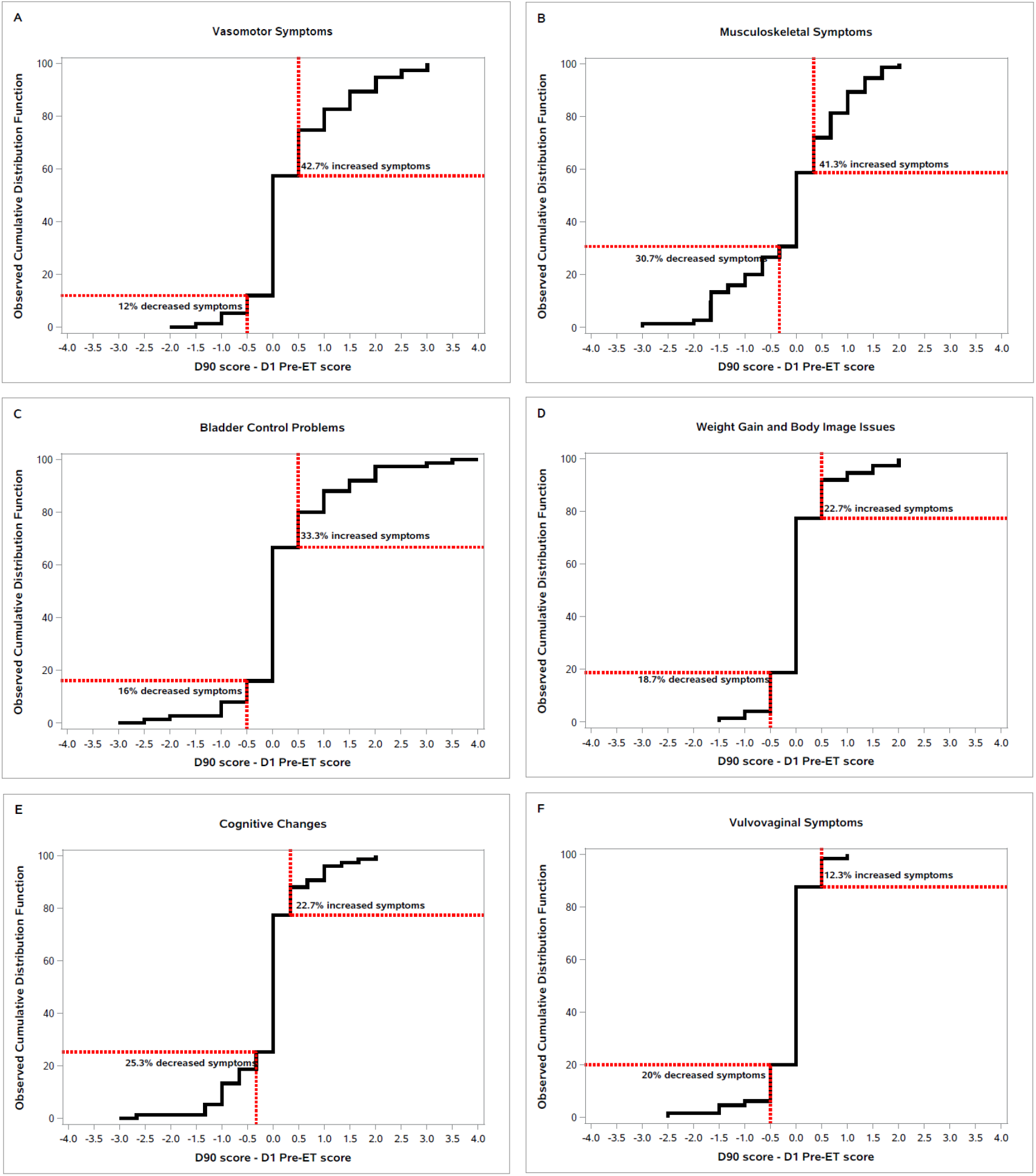
Change in breast cancer symptom burden from baseline to day 90 of Pre-ET. Symptom clusters (A-F) assessed by Breast Cancer Prevention Trial Symptom Checklist (BCPT-SCL) at baseline and at day 90 of Pre-ET. BCPT-SCL symptom cluster scores range from 0-4, with a positive change indicating worsening from baseline. The X-axis represents the difference between Pre-ET day 1 and day 90 scores, and the Y-axis shows the cumulative percentage of patients with at least that score difference.

### Early Discontinuation of Treatment

Eight patients stopped pre-ET early with a median time until discontinuation of 35 days (min: 14 days, max: 64 days). In this group, five participants took AIs and three took tamoxifen. Seven participants discontinued treatment due to intolerable side effects or adverse events (Table 2). One participant stopped pre-ET early citing anxiety about delaying surgery as the reason for discontinuation.

**Table 2.**
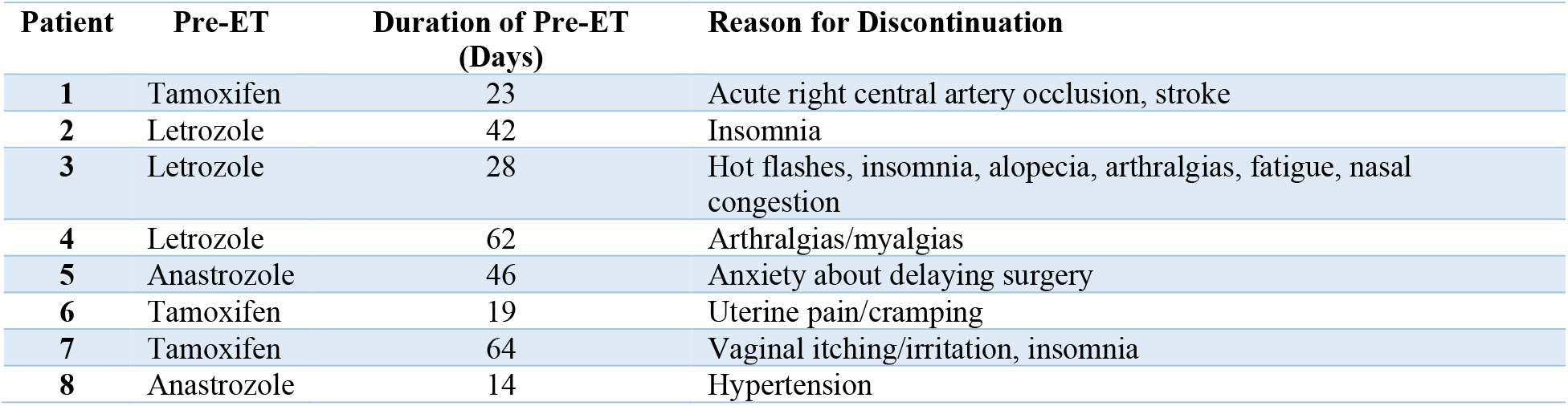
Early Discontinuation of Pre-ET.

### Perceived Sensitivity to Medicine

At baseline, 16% of participants had high PSM scores (≥ 17). High patient-reported PSM was not associated with a worsening HRQoL during pre-ET, as 33% of patients with high PSM and 33% with low PSM reported worsening HRQoL during pre-ET. Participants with high PSM more often reported worsening depression compared to those with low PSM (33.3% versus 12.7%). For most symptom clusters measured by BCPT-SCL, patients with high baseline PSM had a greater increase in symptom burden when taking pre-ET than those with low baseline PSM (Table 3). The opposite was true for worsening vulvovaginal symptoms, which were reported in 8.3% of the high PSM group and 11.1% in the low group (Figure 2).

**Table 3.** Patients with Increased Symptom Burden during Pre-ET compared by PSM.

|  | **High PSM**<br>**n = 12** | **Low PSM**<br>**n = 63** |
| --- | --- | --- |
| Vasomotor Symptoms | 75.0% (9) | 36.5% (23) |
| Musculoskeletal Symptoms | 75.0% (9) | 34.9% (22) |
| Cognitive Changes | 50.0% (6) | 17.5% (11) |
| Weight Gain and Body Image Issues | 41.7% (5) | 19.1% (12) |
| Bladder Control Problems | 41.7% (5) | 31.8% (20) |
| Vulvovaginal Symptoms | 8.3% (1) | 11.1% (7) |

**Figure 2:**
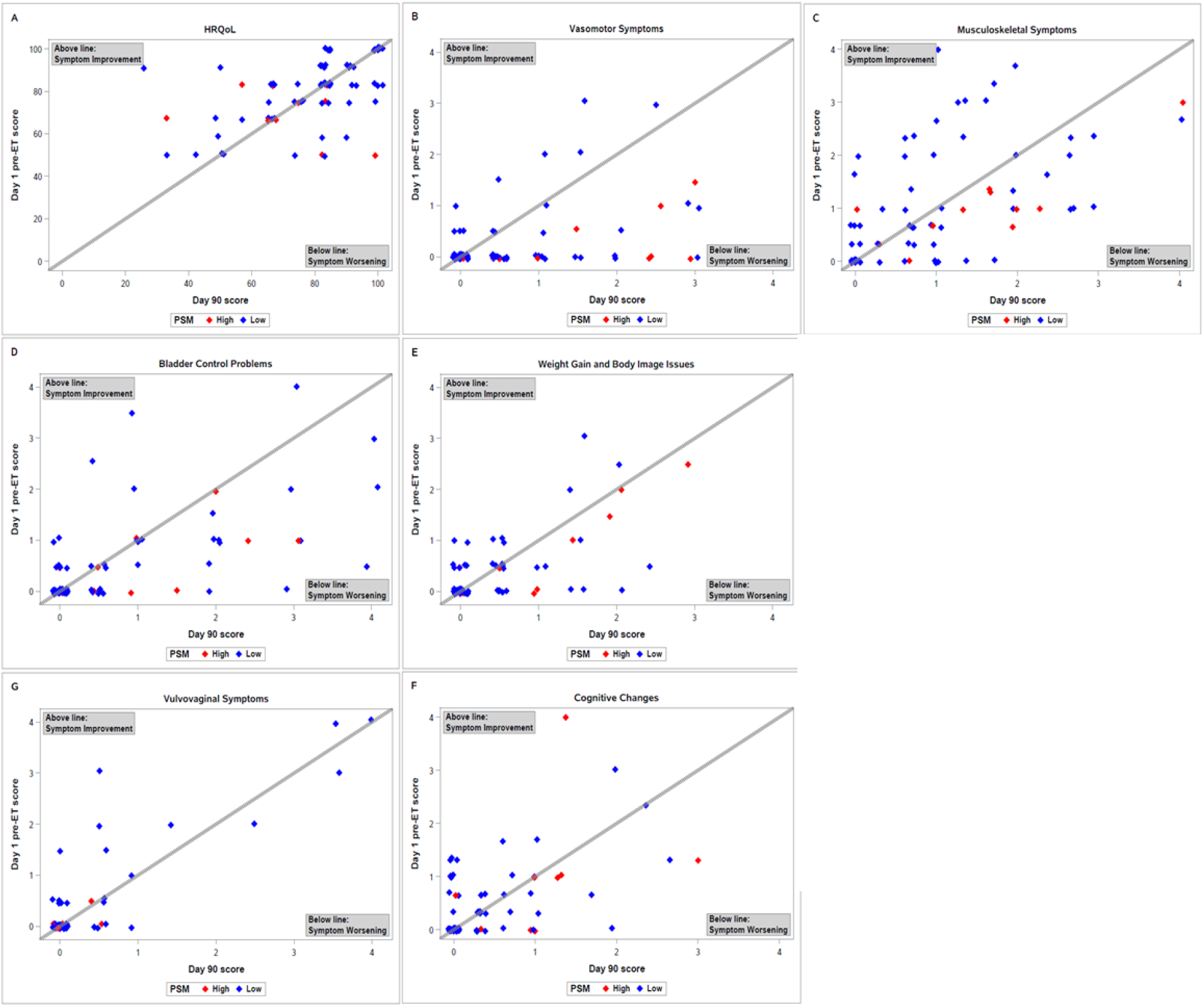
Change in PROs from baseline to Day 90 by Perceived Sensitivity to Medicine (PSM) HRQoL (A) was measured by EORTC-QLQ C30 with a score range of 0 to 100 with a higher score representing a higher/improved functional level. Symptom clusters (B-G) were measured by BCPT-SCL with a range of 0-4 with higher scores indicating worse symptoms. The plotted Day 1 and Day 90 symptom scores show an individual patient's change in symptom severity. Patient data points are color-coded to show those with a high perceived sensitivity to medicine (PSM) and those with a low PSM. HRQoL (A) did not differ by PSM. Patients with high PSM had worsening symptoms for multiple clusters (B-F). but not for vulvovaginal symptoms (G).

## Discussion

Participants in the POWER trial were treated with 90 days of pre-ET to inform patients and physicians about an individual’s ET tolerance and potentially guide adjuvant therapy decisions. The present study evaluated the PROs of POWER trial participants before and after 90 days of pre-ET. Similar to AET, patients’ tolerance of pre-ET and symptoms experienced with pre-ET varied. Most patients (73.3%) developed at least one typical symptom during pre-ET, with vasomotor instability and musculoskeletal symptoms being the most common. A small subset of patients (10.7%) experienced symptoms that resulted in early discontinuation of pre-ET. Most patients reported preserved HRQoL. Ninety days of pre-ET provided adequate exposure for patients to gain individualized knowledge about potential AET side effects and their ability to tolerate them.

The CALGB 9343 and PRIME II trials provided prospective randomized evidence that RT omission does not result in worse overall survival for older women with node-negative ER+ breast cancer with tumors up to 3 cm.^1,2^ These trials established RT omission as an appropriate treatment approach for this population. With this change in the treatment paradigm, adherence to AET for at least 5 years is especially important to ensure an acceptably low recurrence rate in patients who forego radiation. However, poor adherence to AET is a frequent problem that is difficult to predict on an individual patient level. In the POWER trial, 90 days of pre-ET was used as an early test of ET tolerance to identify patients at risk of non-adherence who may be more appropriately treated with RT.

In the ELPh trial, a prospective randomized trial comparing letrozole and exemestane, early change in PROs, notably musculoskeletal side effects, occurred within 3 months of starting AET and was associated with non-adherence to AET at 2 years.^15^ Worsening HRQoL, musculoskeletal symptoms, mood, cognitive function, and weight/body image were associated with increased risk of early cessation of AET. In the MA.27 trial, a randomized controlled trial designed to compare the safety and efficacy of anastrozole and exemestane, AET side effects measured by PROs also developed within 3 months of starting ET.^10^ This study found an association between increased joint pain severity in the first 3 months and the rate of early AET discontinuation. The ELPh and MA.27 PRO provided the scientific rationale for using 90 days as the pre-ET treatment length. Since the majority of the POWER trial participants experienced at least one side effect during pre-ET, this current analysis supports 90 days as an adequate amount of time for patients to take ET and make a more informed decision about their potential tolerance of AET.

Not only did most patients in the POWER trial experience at least one symptom during pre-ET, but the specific symptoms mirrored typical ET symptoms seen in the adjuvant setting. Large prospective AET trials provide the framework for the common side effects of ET, captured as adverse events (AEs) or PROs, with the most commonly cited being hot flashes (13 to 47%) and musculoskeletal symptoms (4 to 47%).^16–23^ Similarly, during the 90 days of pre-ET, 41.3% of patients experienced musculoskeletal symptoms and 42.7% experienced vasomotor symptoms.

As demonstrated in both the ELPH and MA.27 trials, these early ET symptoms may signal an increased risk of long-term non-adherence to AET.^8,10^ Given that symptoms experienced with pre-ET are similar to those reported in the adjuvant setting, we hypothesize that PROs in the pre-ET period will predict a patient’s long-term experience with AET and possible premature discontinuation of AET.

To further understand patients’ ability to tolerate medications, POWER trial participants’ baseline self-reported belief about their medication sensitivity was assessed and compared to symptoms reported during the pre-ET period. In the MA.27 trial, patients who described themselves as more bothered by medication side effects were more likely to discontinue AET early before completing the prescribed course.^10^ Only 16% of POWER trial participants considered themselves highly sensitive to medications, which limited our analysis. However, patients with high baseline PSM did have increased rates of worsening symptom burden for vasomotor symptoms, musculoskeletal symptoms, weight and body image issues, cognitive changes, bladder control problems, and depression when compared to those with low baseline PSM. While most patients in the POWER trial did not have a baseline high PSM, those who did reported more side effects attributed to pre-ET and thus may be at risk for future nonadherence to AET. PROs, such as the PSM survey, should be considered when patients and physicians make adjuvant therapy recommendations and decisions. Future analyses will examine the relationship of baseline PSM to long-term AET adherence for POWER trial participants.

The current study has limitations. Specifically, since the POWER trial was powered to evaluate the primary endpoint, the impact of pre-ET on patient and physician preference towards RT, the statistical analysis for PRO data is limited. Despite this, the current study successfully evaluated changes in PROs before and after pre-ET. Our analysis of PSM was limited by the small number of participants who considered themselves highly sensitive to medications. Another current limitation is the lack of long-term AET adherence data. As discussed, prior studies demonstrate that symptoms experienced in the first 90 days of AET predict potential nonadherence to AET. Longer follow-up of the POWER trial patients will allow for an analysis of how pre-ET tolerance relates to AET adherence, which will be analyzed in future studies.

## Conclusion

In the POWER trial, patients treated with 90 days of pre-ET developed symptoms similar to those experienced with AET. As seen in previously published studies and this current study, 90 days is a sufficient amount of exposure to ET for patients to develop side effects. Future analyses will solidify the correlation between PROs during pre-ET and long-term AET adherence. Assessing symptoms in the pre-operative period provides individualized information to better estimate AET tolerance and may ultimately be used to tailor adjuvant therapy plans to each patient.

## Data Availability

All data produced in the present study are available upon reasonable request to the authors.

## References

1. Hughes KS, Schnaper LA, Bellon JR, et al. Lumpectomy plus tamoxifen with or without irradiation in women age 70 years or older with early breast cancer: long-term follow-up of CALGB 9343. J Clin Oncol Off J Am Soc Clin Oncol. 2013;31(19):2382–2387. doi:10.1200/JCO.2012.45.2615

2. Kunkler IH, Williams LJ, Jack WJL, Cameron DA, Dixon JM, PRIME II investigators. Breast-conserving surgery with or without irradiation in women aged 65 years or older with early breast cancer (PRIME II): a randomised controlled trial. Lancet Oncol. 2015;16(3):266–273. doi:10.1016/S1470-2045(14)71221-5

3. National Comprehensive Cancer Network. Breast Cancer (Version 6.2024). Accessed January 21, 2025. https://www.nccn.org/professionals/physician_gls/pdf/breast.pdf

4. Chu QD, Zhou M, Medeiros KL, Peddi P, Wu XC. Impact of CALGB 9343 Trial and Sociodemographic Variation on Patterns of Adjuvant Radiation Therapy Practice for Elderly Women (≥70 Years) with Stage I, Estrogen Receptor-positive Breast Cancer: Analysis of the National Cancer Data Base. Anticancer Res. 2017;37(10):5585–5594. doi:10.21873/anticanres.11992

5. Keim-Malpass J, Anderson RT, Balkrishnan R, Desai RP, Showalter SL. Evaluating the Long-Term Impact of a Cooperative Group Trial on Radiation Use and Adjuvant Endocrine Therapy Adherence Among Older Women. Ann Surg Oncol. 2020;27(9):3458–3465. doi:10.1245/s10434-020-08430-9

6. Murphy CC, Bartholomew LK, Carpentier MY, Bluethmann SM, Vernon SW. Adherence to adjuvant hormonal therapy among breast cancer survivors in clinical practice: a systematic review. Breast Cancer Res Treat. 2012;134(2):459–478. doi:10.1007/s10549-012-2114-5

7. Chlebowski RT, Geller ML. Adherence to endocrine therapy for breast cancer. Oncology. 2006;71(1-2):1–9. doi:10.1159/000100444

8. Henry NL, Azzouz F, Desta Z, et al. Predictors of aromatase inhibitor discontinuation as a result of treatment-emergent symptoms in early-stage breast cancer. J Clin Oncol Off J Am Soc Clin Oncol. 2012;30(9):936–942. doi:10.1200/JCO.2011.38.0261

9. Meneveau MO, Keim-Malpass J, Camacho TF, Anderson RT, Showalter SL. Predicting adjuvant endocrine therapy initiation and adherence among older women with early-stage breast cancer. Breast Cancer Res Treat. 2020;184(3):805–816. doi:10.1007/s10549-020-05908-8

10. Wagner LI, Zhao F, Goss PE, et al. Patient-reported predictors of early treatment discontinuation: treatment-related symptoms and health-related quality of life among postmenopausal women with primary breast cancer randomized to anastrozole or exemestane on NCIC Clinical Trials Group (CCTG) MA.27 (E1Z03). Breast Cancer Res Treat. 2018;169(3):537–548. doi:10.1007/s10549-018-4713-2

11. Aaronson NK, Ahmedzai S, Bergman B, et al. The European Organization for Research and Treatment of Cancer QLQ-C30: a quality-of-life instrument for use in international clinical trials in oncology. J Natl Cancer Inst. 1993;85(5):365–376. doi:10.1093/jnci/85.5.365

12. Radloff LS. The CES-D Scale: A Self-Report Depression Scale for Research in the General Population. Appl Psychol Meas. 1977;1(3):385–401. doi:10.1177/014662167700100306

13. Stanton AL, Bernaards CA, Ganz PA. The BCPT symptom scales: a measure of physical symptoms for women diagnosed with or at risk for breast cancer. J Natl Cancer Inst. 2005;97(6):448–456. doi:10.1093/jnci/dji069

14. Horne R, Faasse K, Cooper V, et al. The perceived sensitivity to medicines (PSM) scale: an evaluation of validity and reliability. Br J Health Psychol. 2013;18(1):18–30. doi:10.1111/j.2044-8287.2012.02071.x

15. Kadakia KC, Snyder CF, Kidwell KM, et al. Patient-Reported Outcomes and Early Discontinuation in Aromatase Inhibitor-Treated Postmenopausal Women With Early Stage Breast Cancer. The Oncologist. 2016;21(5):539–546. doi:10.1634/theoncologist.2015-0349

16. Baum M, Budzar AU, Cuzick J, et al. Anastrozole alone or in combination with tamoxifen versus tamoxifen alone for adjuvant treatment of postmenopausal women with early breast cancer: first results of the ATAC randomised trial. Lancet Lond Engl. 2002;359(9324):2131–2139. doi:10.1016/s0140-6736(02)09088-8

17. Breast International Group (BIG) 1-98 Collaborative Group, Thürlimann B, Keshaviah A, et al. A comparison of letrozole and tamoxifen in postmenopausal women with early breast cancer. N Engl J Med. 2005;353(26):2747–2757. doi:10.1056/NEJMoa052258

18. Goss PE, Ingle JN, Pritchard KI, et al. Exemestane versus anastrozole in postmenopausal women with early breast cancer: NCIC CTG MA.27--a randomized controlled phase III trial. J Clin Oncol Off J Am Soc Clin Oncol. 2013;31(11):1398–1404. doi:10.1200/JCO.2012.44.7805

19. Boccardo F, Rubagotti A, Puntoni M, et al. Switching to anastrozole versus continued tamoxifen treatment of early breast cancer: preliminary results of the Italian Tamoxifen Anastrozole Trial. J Clin Oncol Off J Am Soc Clin Oncol. 2005;23(22):5138–5147. doi:10.1200/JCO.2005.04.120

20. Coombes RC, Kilburn LS, Snowdon CF, et al. Survival and safety of exemestane versus tamoxifen after 2-3 years’ tamoxifen treatment (Intergroup Exemestane Study): a randomised controlled trial. Lancet Lond Engl. 2007;369(9561):559–570. doi:10.1016/S0140-6736(07)60200-1

21. Burstein HJ. Aromatase inhibitor-associated arthralgia syndrome. Breast Edinb Scotl. 2007;16(3):223–234. doi:10.1016/j.breast.2007.01.011

22. Crew KD, Greenlee H, Capodice J, et al. Prevalence of joint symptoms in postmenopausal women taking aromatase inhibitors for early-stage breast cancer. J Clin Oncol Off J Am Soc Clin Oncol. 2007;25(25):3877–3883. doi:10.1200/JCO.2007.10.7573

23. Henry NL, Giles JT, Ang D, et al. Prospective characterization of musculoskeletal symptoms in early stage breast cancer patients treated with aromatase inhibitors. Breast Cancer Res Treat. 2008;111(2):365–372. doi:10.1007/s10549-007-9774-6

